# Low plasma 25(OH) vitamin D level is associated with increased risk of COVID-19 infection: an Israeli population-based study

**DOI:** 10.1101/2020.07.01.20144329

**Authors:** Eugene Merzon, Dmitry Tworowski, Alessandro Gorohovski, Shlomo Vinker, Avivit Golan Cohen, Ilan Green, Milana Frenkel Morgenstern

## Abstract

**Aim:** To evaluate associations of plasma 25(OH)D status with the likelihood of coronavirus disease (COVID-19) infection and hospitalization.

**Methods:** The study population included the 14,000 members of Leumit Health Services who were tested for COVID-19 infection from February 1^st^ to April 30^th^ 2020, and who had at least one previous blood test for plasma 25(OH)D level. “Suboptimal” or “low” plasma 25(OH)D level was defined as plasma 25-hydroxyvitamin D, or 25(OH)D, concentration below 30 ng/mL.

**Results:** Of 7,807 individuals, 782 (10.1%) were COVID-19-positive, and 7,025 (89.9%) COVID-19-negative. The mean plasma vitamin D level was significantly lower among those who tested positive than negative for COVID-19 [19.00 ng/mL (95% confidence interval [CI] 18.41-19.59) *vs*. 20.55 (95% CI 20.32-20.78)]. Univariate analysis demonstrated an association between low plasma 25(OH)D level and increased likelihood of COVID-19 infection [crude odds ratio (OR) of 1.58 (95% CI 1.24-2.01, p<0.001)], and of hospitalization due to the SARS-CoV-2 virus [crude OR of 2.09 (95% CI 1.01-4.30, p<0.05)]. In multivariate analyses that controlled for demographic variables, and psychiatric and somatic disorders, the adjusted OR of COVID-19 infection [1.45 (95% CI 1.08-1.95, p<0.001)], and of hospitalization due to the SARS-CoV-2 virus [1.95 (95% CI 0.98-4.845, p=0.061)] were preserved. In the multivariate analyses, age over 50 years, male gender and low-medium socioeconomic status were also positively associated with the risk of COVID-19 infection; age over 50 years was positively associated with the likelihood of hospitalization due to COVID-19.

**Conclusion:** Low plasma 25(OH)D level appears to be an independent risk factor for COVID-19 infection and hospitalization.

## INTRODUCTION

From its origin in Wuhan, China in December 2019, the novel coronavirus disease, COVID-19, caused by severe acute respiratory syndrome coronavirus 2 (SARS-CoV-2) virus, has spread rapidly throughout the world (1). In Israel, the first case of the COVID-19 infection was reported on February 21, 2020. On March 11, 2020 the World Health Organization (WHO) declared COVID-19 disease a global pandemic (2). Immediate targeted action is needed to identify risk factors of COVID-19. The SARS-CoV-2 virus has high levels of transmissibility, estimated basic reproduction (R_o_) ranging from 2.6 to 4.7, and an average incubation duration ranging from 2 to 14 days (3). The main routes of transmission are respiratory droplets and direct contact with contaminated objects and surfaces (4).

The status of the immune system is determined by a multitude of factors that may contribute to the risk of a viral infection (5-16). Vitamin D is recognized as an important co-factor in several physiological processes linked with bone and calcium metabolism, and also in diverse non-skeletal outcomes, including autoimmune diseases, cardiovascular diseases, diabetes type 2, obesity and cognitive decline and infections (17,18). In particular, the pronounced impact of vitamin D metabolites on the immune system response, and on the development of COVID-19 infection by the novel SARS CoV-2 virus, has been described (5-16). Vitamin D deficiency has been recognized as a worldwide pandemic (19,20). We aimed to determine associations of low plasma 25(OH)D with the risk of COVID-19 infection and hospitalization, using real-world data. We hypothesized that the mean plasma level of 25(OH)D would be significantly lower and, accordingly, the rate of suboptimal plasma 25(OH)D would be significantly higher, among persons testing positive for COVID-19 infection, and among persons subsequently hospitalized, in a large population-based data sample.

## METHODS

We conducted a population-based study utilizing data from the Leumit Health Services (LHS) database, a large health maintenance organization in Israel that provides services to around 730,000 members nationwide. The comprehensive computerized database of LHS is continuously updated with regard to demographics, medical visits, laboratory tests and hospitalizations. The validity of the diagnoses in the registry is high for important medical diagnoses and laboratory data (21-23). The study period was from February 1^st^ to April 30^th^ 2020. The study population included all members of LHS who were tested for COVID-19 infection during the study period and who had at least one previous test for plasma 25(OH)D level (7,807 subjects). Referrals for viral tests were according to Israeli Ministry of Health guidelines (March 2020). COVID-19 testing was done only by physician referral (based on clinical criteria of exposure to confirmed COVID-19 patients or symptoms suggesting COVID-19) using the AllplexTM 2019-nCoV Assay (Seegene Inc., Seoul, Republic of Korea) (24). According to LHS guidelines, blood was collected from fasting persons and transported on ice to the Center Laboratory for processing within 4 h of collection using DiaSorin Chemiluminescence assay (25-28). Data of each subject were collected from the LHS computerized database and included age, gender, socioeconomic status (SES), weight, height, body mass index, current smoking status, psychiatric and somatic comorbidities, and hospitalizations as a result of the COVID-19 infection.

### Definitions

All the somatic and psychiatric diagnoses were based on the International Classification of Disease, tenth revision (ICD-10) codes and included chronic lung disorders (asthma, chronic obstructive pulmonary disease), diabetes, hypertension, depressive and anxiety disorders, schizophrenia and dementia.

***SES*** was defined according to a person’s home address. The Israeli Central Bureau of Statistics classifies all cities and towns into 20 subgroups of SES. The classifications of one to nine were considered as a low-medium SES, and ten to twenty were considered as medium-high SES.

***Obesity*** was considered as BMI>30 m^2^/kg.

According to Endocrine Society, National Osteoporosis Foundation, and International Osteoporosis Foundation, the optimal 25(OH) D levels should be greater than 30 ng/mL (75 nmol/L), thus the plasma 25(OH)D level that is less than 30 ng/mL (75 nmol/L) were considered as ***suboptimal and referred as “low” in our study*** *(29-31) (see Supplementary Table S1)*.

### Statistical analysis

Statistical analysis was conducted using STATA 12 software (StataCorp LP, College Station, TX). The initial analysis compared demographic characteristics between individuals who tested positive (COVID-19-P) and negative (COVID-19-N) for COVID-19. Student’s t-test and Fischer’s exact χ^2^ test were used for continuous and categorical variables, respectively, based on a normal distribution (0,1) and variable characteristics. The categorical data were shown in counts and percentages. Data on continuous variables with normal distribution were presented as means and 95% confidence intervals (CIs). The assumptions were based on two-sided tests with *α of 0*.*05*.

Preliminary evaluation of risk estimates was conducted by stratified analyses. Subsequently, multivariate logistic regression was used to estimate the odds ratios (OR) and 95% CI for the independent association between low plasma 25(OH)D and a positive PCR test for the SARS-CoV-2 virus, while controlling for potential confounders. The association of low plasma 25(OH)D level with hospitalization due to COVID-19 infection was assessed among those who tested positively for COVID-19.

### Visualization methods

Open source programs, particularly, Plotly R Open Source Graphing Library was used. Plotly’s R graphing library was used for the production of figures, including scatter plots, area charts, bar charts, and 3D charts.

## RESULTS

### Low vitamin D level and the likelihood of COVID-19 infection

Of 14,022 subjects, aged 2 months to 103 years, who were tested for COVID-19 infection, 1,416 (10.1%) had at least one positive result; and 12,606 (89.9%) had only negative results (Figure 1). After excluding the 6,215 individuals without data on plasma 25(OH)D levels, the study sample composed 7,807 (Figure 1). Also for this sample, the proportion of infected individuals was 10.1% (782/7,807) for COVID-19-P, and 7,025 (89.9%) for COVID-19-N. In a primary univariate analysis, COVID-19-P subjects were younger, and more likely to be males and to reside in a lower SES area than were COVID-19-N subjects (Table 1, Figures S1A and S1B, Supplementary Material). The mean plasma 25(OH)D level was significantly lower for COVID-19-P subjects (Table 1), and the proportion with low vitamin D level was higher (89.90% vs. 84.91%, p<0.001) (Table 2). The prevalence of dementia, hypertension, cardiovascular disease and chronic lung disorders were greater among persons who were COVID-19-N (Figure S1A, Supplementary Material) than those who were COVID-19-P (Figure S1B, Supplementary Material) (p<0.05, p<0.001, p<0.001, p<0.001) (Table 2).

**Table 1:** Demographic characteristics of the study sample stratified by COVID-19 test results.

| Demographics | Covid-19 - positive<br>n= 782 (10.1%) | Covid-19 - negative<br>n= 7,025 (89.9%) | P-value |
| --- | --- | --- | --- |
| Mean age, (years, 95%CI) | 35.58 (34.49; 36.67) | 47.35 (46.87-47.85) | 0.001 |
| Age categories N (%) |  |  |  |
| 0-5 years | 3(0.38%) | 18(0.26%) | 0.023 |
| 5-20 years | 79(10.10%) | 381(5.42%) | 0.001 |
| 20-40 years | 249(31.84%) | 2,504(35.64%) | 0.036 |
| 40-60 years | 266(34.02%) | 2,082(29.64%) | 0.001 |
| 60-80 years | 152(19.44%) | 1,378(19.62%) | 0.082 |
| 80+ years | 33(4.22%) | 662(9.42%) | 0.001 |
| SES * |  |  |  |
| Low-medium | 601(83.70%) | 4,418(67.73%) | 0.001 |
| High-medium | 117(16.30%) | 2,105(32.27%) | 0.001 |
| Gender N (%) |  |  |  |
| **Male** | **385(49.23%)** | **2,849(40.56%)** | **0.001** |
| Female | 397(50.77%) | 4,176(59.44%) | 0.001 |
| Smoking N (%) | 127(18.70%) | 1,136(19.39%) | 0.056 |
| Mean BMI, (95%CI) | 27.32( 26.88-27.77) | 27.36(27.22-27.52) | 0.432 |
| Mean vitamin D(ng/mL; 95%CI) | 19.00(18.41-19.59) | 20.55(20.32-20.78) | 0.026 |

*Note.* \*SES – residential socioeconomic status;

**Table 2:** Clinical characteristics of the study sample stratified by COVID-19 test results.

| **Variable N (%)** | **Covid-19 - positive**<br>**n= 782 (10.1%)** | **Covid-19 - negative**<br>**n= 7,025 (89.9%)** | **P-value** |
| --- | --- | --- | --- |
| Low vitamin D level** | **703(89.90%)** | **5,965(84.91%)** | **0.001** |
| Smoking | 127(18.70%) | 1,136(19.39%) | 0.669 |
| Depression/Anxiety | 73(9.34%) | 817(11.63%) | 0.055 |
| Schizophrenia | 15(1.92%) | 141(2.01%) | 0.866 |
| Dementia | **27(3.45%)** | **427(6.08%)** | **0.025** |
| Diabetes mellitus | 154(19.69%) | 1,578(22.46%) | 0.055 |
| Hypertension | **174(22.25%)** | **1,962(27.93%)** | **0.046** |
| Cardiovascular disease | **78(9.97%)** | **1,172(16.68%)** | **0.001** |
| Chronic lung disorders | **66(8.44%)** | **935(13.31%)** | **0.001** |
| Obesity | 235(33.05%) | 1,900(29.66%) | 0.350 |

Note: \*\* **Low plasma 25(OH)D level - the total plasma levels less than 30 ng/mL.**

**Figure 1.**
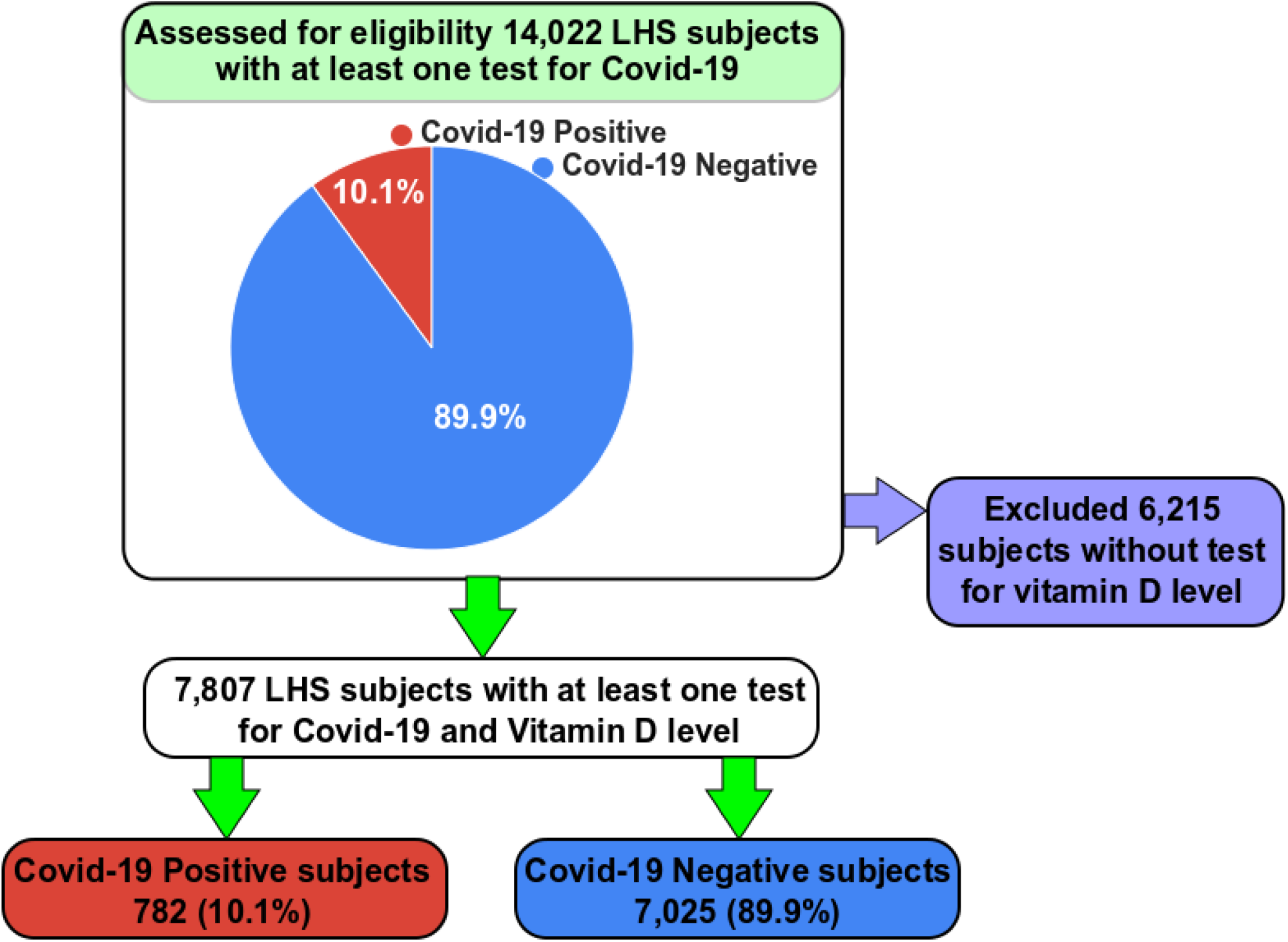
Flowchart of the study design.

Multivariate analysis, after controlling for the demographic variables, and psychiatric and somatic disorders demonstrated an independent and significant association between a low 25(OH)D level and the increased likelihood of COVID-19 infection [adjusted OR of 1.50 (95% CI 1.13-1.98, p<0.001)]. The risk of COVID-19 infection was independently positively associated with being male (Figure S1B, Supplementary Material) [adjusted OR of 1.49 (95% CI 1.24-1.79, p<0.05)], aged older than 50 years (Figures S1A and S1B, Supplementary Material) [adjusted OR of 1.56 (95% CI 1.26-1.92, p<0.05)], and residing in a low-medium SES city or town (Figure S1B, Supplementary Material) [adjusted OR of 2.06 (95% CI 1.65-2.59, p<0.001)]. Independent negative associations were observed between the risk of COVID-19 infection and having a diagnosis of dementia [adjusted OR of 0.56 (95%CI 0.32-0.98, p<0.05], of cardiovascular disease [adjusted OR of 0.59 (95%CI 0.44-0.79 p<0.001] and of a chronic lung disorder [adjusted OR of 0.58 (95%CI 0.42-0.79 p<0.001] (Table 3, Figures S1A and S1B, Supplementary Material)).

**Table 3:** Multivariate logistic regression analysis of the odds ratio for infection with COVID-19, controlling for multiple conditions.

| **Variable** | **Crude OR (95% CI)** | **P value** | **Adjusted OR (95% CI)** | **P value** |
| --- | --- | --- | --- | --- |
| Low vitamin D level** | 1.58(1.24- 2.01) | **0.001** | 1.50(1.13-1.98) | **0.001** |
| Age over 50 years | 1.51(1.21- 1.89) | **0.001** | 1.56(1.26- 1.92) | **0.001** |
| Male | 1.42(1.23-1.65) | **0.001** | 1.49(1.24-1.79) | **0.001** |
| Low-medium- SES | 2.45(1.99-3.01) | **0.001** | 2.13(1.69- 2.68) | **0.001** |
| Smoking | 0.95(0.78- 1.17) | 0.669 | **0.71(0.56-0 .91)** | **0.05** |
| Depression/Anxiety | 0.78(0.61-1.01) | 0.062 | 1.13(0.84-1.51) | 0.423 |
| Schizophrenia | 0.95(0.56-1.63) | 0.478 | 1.01(0.54-1.86) | 0.991 |
| Dementia | 0.55(0.29- 0.84) | **0.001** | **0.56(0.32- 0.98)** | **0.006** |
| Diabetes mellitus | 0.84(0.71-1.01) | 0.07 | 0.91(0.71-1.17) | 0.469 |
| Hypertension | 0.74(0.62-0.88) | **0.001** | 0.86(0.67-1.11) | 0.670 |
| Cardiovascular disease | 0.55(0.43- 0.71) | **0.001** | **0.58(0.44-0.79)** | **0.001** |
| Chronic lung disorders | 0.60(0.46- 0.78) | **0.001** | **0.58 (0.45-0.76)** | **0.001** |
| **BMI** | **0.99(0.98-1.011)** | **0.857** | **0.99(0.98-1.009)** | **0.523** |

Note: \*\* Low plasma 25(OH)D level Low vitamin D level - the total plasma levels less than 25-(OH)D levels of 30 ng/mL

### Low vitamin D level and the likelihood of hospitalization due to COVID-19 infection

Among the COVID-19-P individuals, those who were hospitalized were older [58.69 years (95%CI 54.78; 62.61) vs. 46.88 (95%CI 46.42-47.35)], and more likely to be male (47.8% vs. 41.3%, p<0.001) and to reside in a city or town of low-medium SES (73.64% vs. 69.45%, p<0.001). The hospitalized compared to non-hospitalized individuals had a significantly lower mean plasma 25-dihydroxy-vitamin D_3_ level [18.38 ng/mL (95% CI 16.79-19.96) vs. 20.45 ng/mL (95% CI 20.22-20.68), p<0.001]. In a univariate analysis, a low plasma 25(OH)D level was associated with an increased likelihood of hospitalization for COVID-19 infection [crude OR of 2.09 (95% CI 1.01-4.31, p<0.05)]. In a multivariate analysis that controlled for demographic variables and chronic disorders, the adjusted OR decreased slightly to 1.95 (95% CI 0.98-4.84, p=0.061). Therefore, in this analysis, only age over 50 years was statistically significant associated with the likelihood for hospitalization due to COVID-19 [adjusted OR of 2.71(95%CI 01.55; 4.78p<0.001] (Table 4 and Figure 3).

**Table 4:** Multivariate logistic regression analysis of the odds ratio for hospitalization of patients with COVID-19, controlling for multiple clinical conditions.

| **Variable** | **Crude OR (95% CI)** | **P value** | **Adjusted OR (95% CI) \*** | **P value** |
| --- | --- | --- | --- | --- |
| Low vitamin D level | 2.09(1.01-4.31) | **0.021** | 1.95(0.99-4.78) | 0.056 |
| **Age over 50 years** | 2.51(1.21-4.89) | **0.001** | **2.71(1.55; 4.78)** | **0.002** |
| Male sex | 1.32(0.76-2.11) | 0.223 | 1.35(0.83- 2.21) | 0.324 |
| Low-medium- SES | 1.24(0.81-1.91) | 0.254 | 1.36(0.83-2.21) | 0.222 |
| Smoking | 1.14(0.68-2.17) | 0.669 | 1.22(0.71-2.08) | 0.470 |
| Depression/Anxiety | 0.78(0.61-1.01) | 0.662 | 0.94(0.50-1.76) | 0.846 |
| Schizophrenia | 0.95(0.56-1.63) | 0.478 | 1.24(0.58-2.67) | 0.581 |
| Dementia | 1.65(0.29- 4.84) | 0.625 | 1.52( 0.46- 4.98) | 0.489 |
| Diabetes mellitus | 2.04(1.39-2.99)) | 0.001 | 1.82(0.41-2.36 ) | 0.696 |
| Hypertension | 1.81(1.49-2.33) | **0.001** | 1.56(0.91-2.71) | 0.113 |
| Cardiovascular disease | 1.54(0.67-3.53) | 0.231 | 1.06(0.44-2.58) | 0.896 |
| Chronic lung disorders | 1.44(0.89-2.34) | 0.142 | 0.94 0.52-1.71) | 0.726 |
| BMI | 1.17(0.98-1.38) | 0.075 | 0.99(0.98-1.011) | p=0.804 |

**Figure 2.**
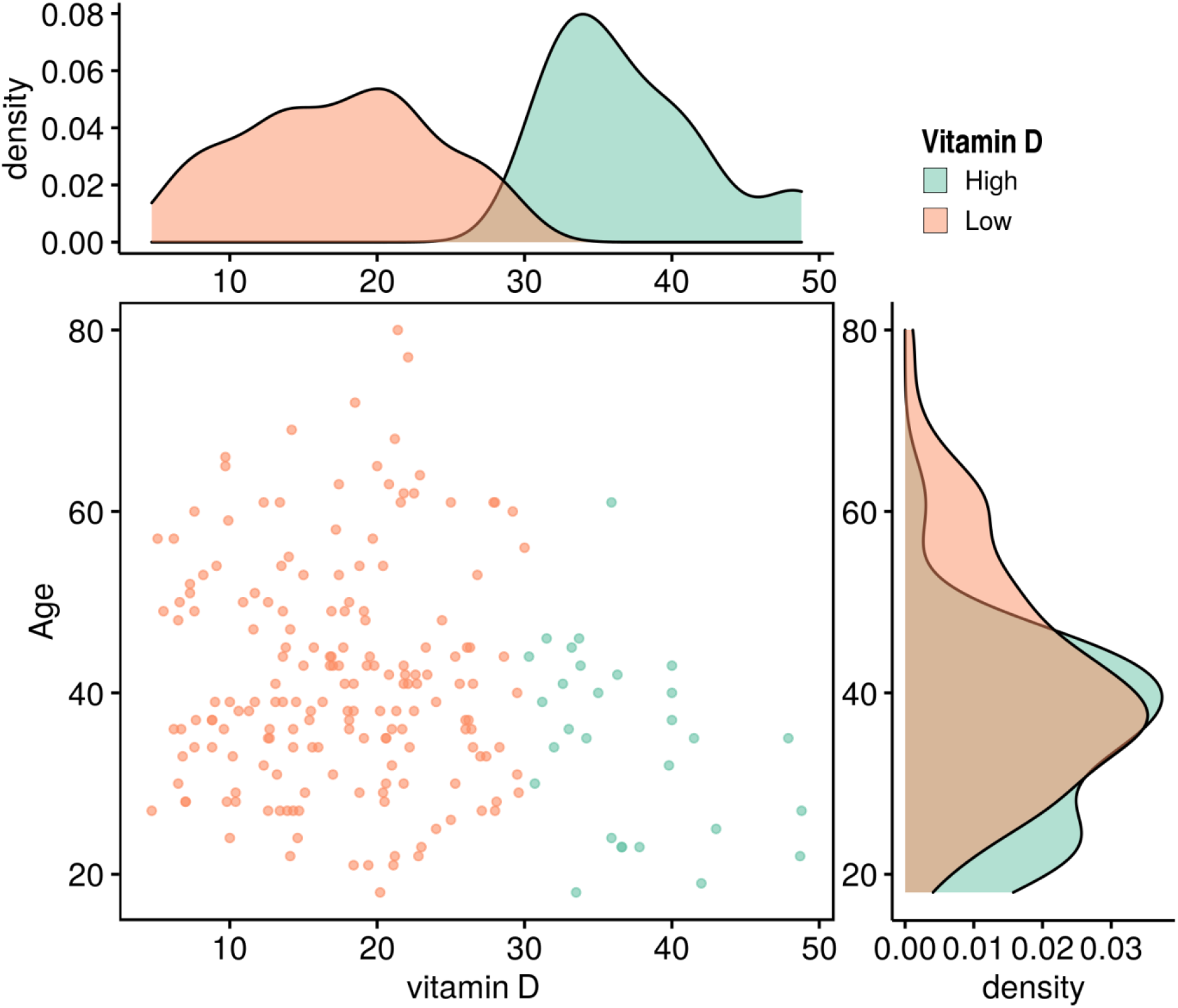
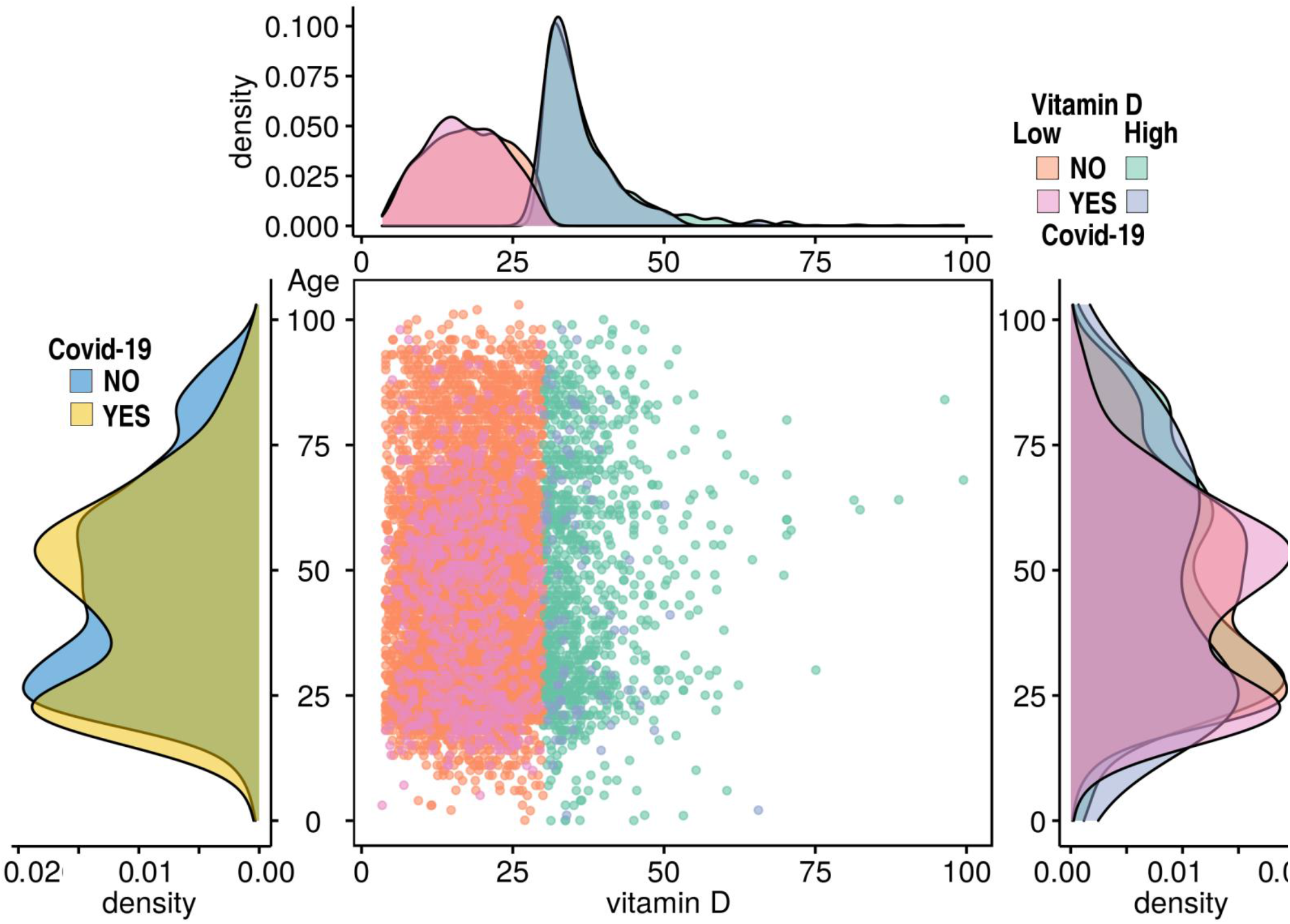
(A) Distribution densities of vitamin D_3_ levels (vertical axis) and age (horizontal axis) among persons infected (A) and not infected (B) with COVID-19. The criterion for vitamin D_3_ deficiency was <30ng/ml.

**Figure 3.**
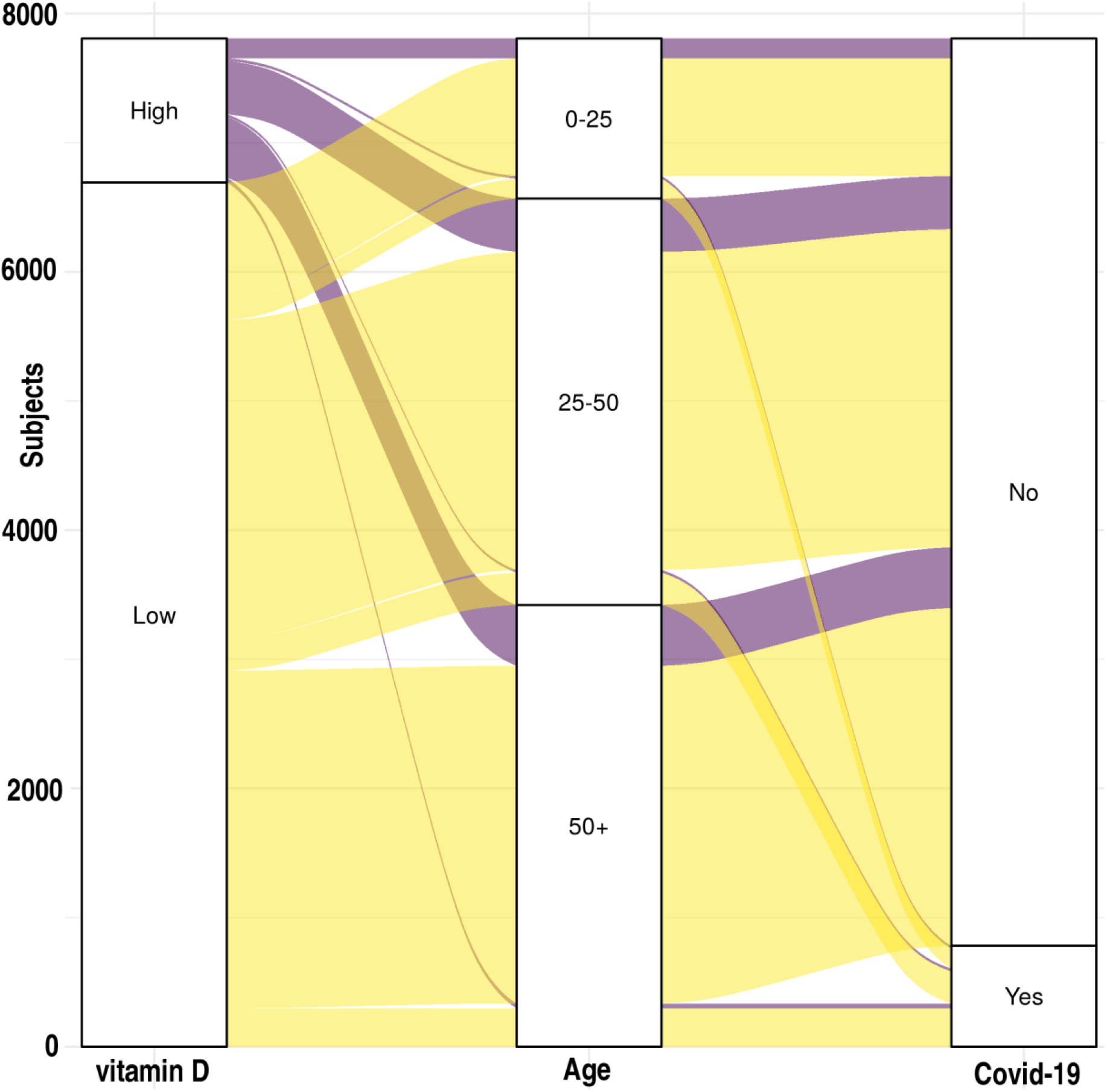
The likelihood of hospitalization due to COVID-19 according to two risk factors: the presence / absence of vitamin D_3_ deficiency and age, classified by: 0-25, 25-50, 50+ years.

## DISCUSSION

The main finding of this study was the low plasma 25(OH)D level association with COVID-19 hospitalization risks, for patients tested positively for COVID-19, after adjusting for age, gender, SES and chronic, mental and physical disorders. Hence, low 25(OH)D level was identified as independently associated with the likelihood of COVID-19 infection. This finding is in agreement with the results of other studies (5,7,9-16,20,32-44). Further, reduced risk of acute respiratory tract infection following vitamin D supplementation has been reported (45,46). Notably, a recent study from the UK (33-35) that included 449 subjects (from the UK Biobank) with confirmed COVID-19 infection did not find an association between vitamin D metabolite concentration and the risk of viral infections (47) as well as COVID-19 infection (5,7,9-16,20,32-44). The discrepancy between those and our results may be explained by a sample size of less than half in that study, the older population and the inability to control for several confounders, like SES and chronic medical conditions.

According to our analysis, persons with COVID-19-P were younger than non-infected ones. Two-peak distributions for age groups were demonstrated to confer increased risk for COVID-19: ages 25 years old and 50 years old (Figure 4). The first peak may be explained by high social gathering habits at the young age. The peak at age 50 years may be explained by continued social habits, in conjunction with various chronic diseases (Figure 4). Other clinical characteristics that were significantly linked to the likelihood of COVID-19 infection included male gender and low residential SES (Figure S1, Supplementary Material). Despite its being discussed as a risk factor in prior publications (48-50), obesity had not been significantly associated with either an increased risk for COVID-19 infection or with hospitalization due to COVID-19 in this study.

**Figure 4.**
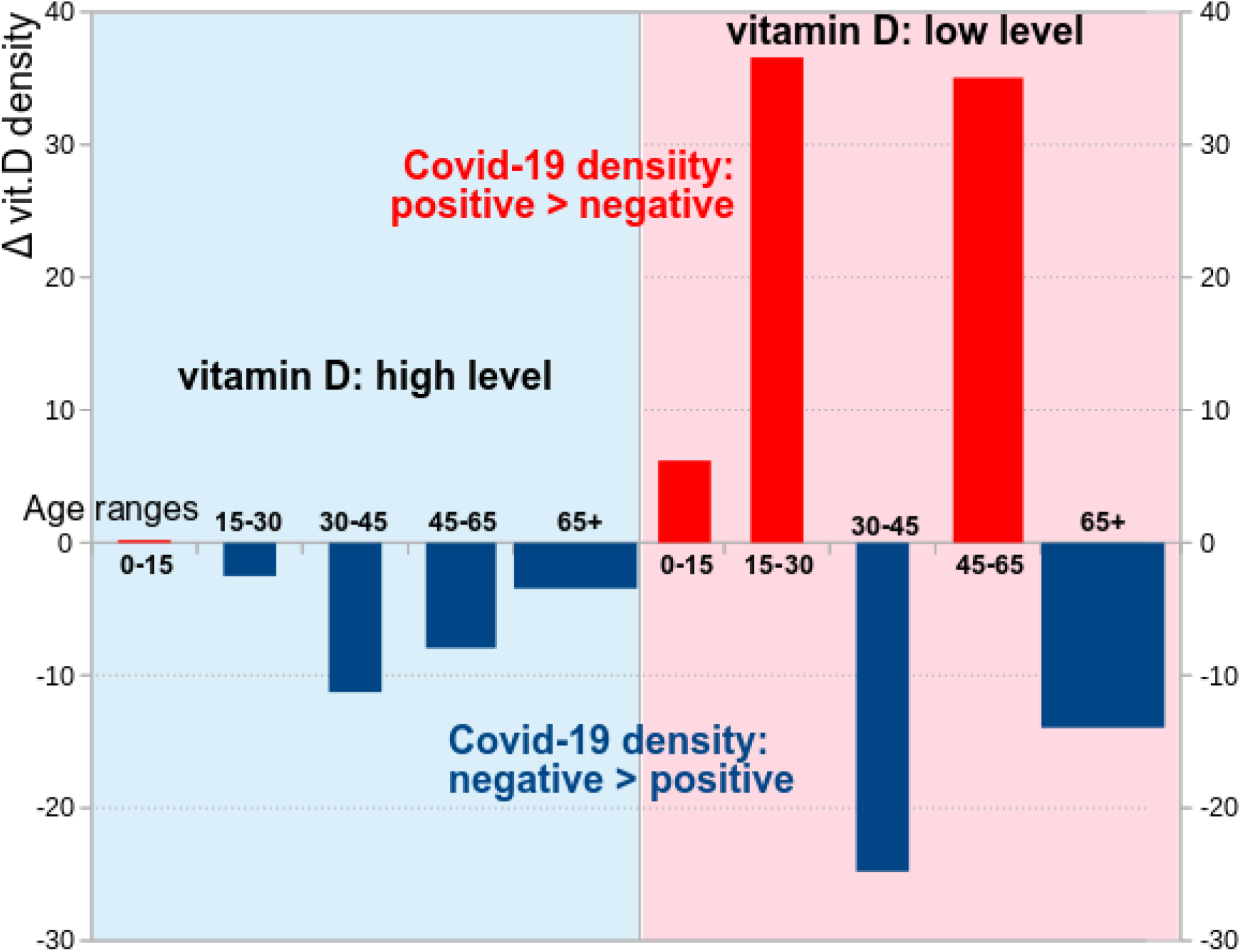
Two-peak age groups as high risk for COVID-19: ages 25 years and 50 years (red bars). Both age groups are included in the subset of vitamin D_3_ deficient patients (the area highlighted pink). In the subset of persons without vitamin D_3_ deficiency, the age range of 30-45 years old peaked (shown in the area highlighted in blue).

Surprisingly, chronic medical conditions, like dementia, cardiovascular disease, and chronic lung disease that were considered to be very risky in previous studies (51,52), were not found as increasing the rate of infection in our study. However, this finding is highly biased by the severe social contacts restrictions that were imposed on all the population and were even more emphasized in this highly vulnerable population. Therefore, we assume, that following **the** Israeli Ministry of Health instructions, patients with chronic medical conditions significantly reduced their social contacts. **This** might indeed minimize their **the** risks of COVID-19 infection **in that group of patients**. The negative association with the current smoking status is unclear and should be investigated, since recent studies show conflicting data (38,53-64). Finally, in a subset analysis of only COVID-19-P subjects, **hospitalized** patients were significantly older (58.69 *vs*. 46.88 years). Multivariate analysis showed that being older than 50 years old was the single statistically significant risk factor for hospitalization. To conclude, the low plasma 25(OH)D levels almost doubled the risk for hospitalization due to the COVID-19 infection in the Israeli studied cohort.

## Conclusions, strengths and limitations

The main strength of the study is its being large, real-world, and population-based. An additional strength is the analysis of a multitude of variables that may affect the risk of COVID-19 infection, independent of plasma 25(OH)D levels. However, the major weakness of the study is the retrospective database design. Data regarding COVID-19 symptoms and the hospitalization due to COVID-19 infection, and also adverse clinical outcomes (for example, mechanical ventilation) will be further assessed. Moreover, a possible selection bias arises in that vitamin D level was tested according to the presentation of symptoms, and not according to population-wide testing. However, our previous study showed that poor health function was not associated with low 25(OH)D levels (47). To conclude, our study found that suboptimal plasma vitamin D levels may be a potential risk factor for COVID-19 infection, particularly, for the high hospitalization risks, independent of demographic characteristics and medical conditions. The finding is important, since it could guide healthcare systems in identifying populations at risk, and contribute to interventions aimed to reduce the risk of the COVID-19 infection. More studies are required, including assessment of the effects of vitamin D_3_ supplements on the risk of hospitalizations due to COVID-19 infection.

## Data Availability

This is a data-based study, and as such, has no clinical trial registration number. The study received approval from the Leumit Health Services research committee and the Shamir Medical Centre IRB.

## Authors’ contribution

EM, MFM, DT, IG, AVG, and SV contributed the research questions, EM and IG performed data mining. EM performed statistical analysis. EM and DT wrote the final draft of the manuscript. MFM, AG, and DT presented results in visual forms, MFM and EM supervised the project, and the project design. All the authors contributed to editing of the manuscript. No honorarium, grant, or other form of payment was given to any of the authors to produce the manuscript.

